# Ivermectin shows clinical benefits in mild to moderate COVID19: A randomised controlled double blind dose response study in Lagos

**DOI:** 10.1101/2021.01.05.21249131

**Authors:** OE Babalola, CO Bode, AA Ajayi, FM Alakaloko, IE Akase, E Otrofanowei, OB Salu, WL Adeyemo, AO Ademuyiwa, S Omilabu

## Abstract

**Introduction:** In vitro studies have shown the efficacy of Ivermectin (IV) to inhibit the SARS - CoV-2 viral replication, but questions remained as to In-vivo applications. We set out to explore the efficacy and safety of Ivermectin in persons infected with COVID19.

**Methods:** We conducted a translational proof of concept (PoC) randomized, double blind placebo controlled, dose response, parallel group study of IV efficacy in RT - PCR proven COVID 19 positive patients. 62 patients were randomized to 3 treatment groups. (A) IV 6mg regime, (B)IV 12 mg regime (given Q84hrs for 2weeks) (C, control) Lopinavir/Ritonavir. All groups plus standard of Care.

**Results:** The Days to COVID negativity [DTN] was significantly and dose dependently reduced by IV (p = 0.0066). The DTN for Control were, = 9.1+/−5.2, for A 6.0 +/− 2.9, and for B 4.6 +/−3.2. 2 Way repeated measures ANOVA of ranked COVID 19 + / − scores at 0, 84, 168, 232 hours showed a significant IV treatment effect (p=0.035) and time effect (p <0.0001). IV also tended to increase SPO2 % compared to controls, p = 0.073, 95% CI - 0.39 to 2.59 and increased platelet count compared to C (p = 0.037) 95%CI 5.55 - 162.55 × 10^3^/ml. The platelet count increase was inversely correlated to DTN (r = −0.52, p = 0.005). No SAE was reported.

**Conclusions:** 12 mg IV regime may have superior efficacy. IV should be considered for use in clinical management of SARS-Cov-2, and may find applications in community prophylaxis in high-risk areas.

## INTRODUCTION

The Corona Virus Disease 2019 (COVID 19) pandemic caused by the Severe Acute Respiratory Syndrome Corona virus −2 (SARS-CoV-2)^1^ led to a World Health Organization declared global pandemic on March 11, 2020^2^. As of December 22, 2020, more than 77.7 million people on all continents had being infected and more than 1.7 million people had died globally^3^. Prolonged morbidity after recovery from acute COVID 19^4^ and astronomical hospital costs^5^, has left hospitals swamped and health workers exhausted. In addition, the global economy is in a depression^6^ with massive job losses, furloughing and social movement restrictions.

There have been concerted attempts to seek preventive and interventional modalities to arrest the spread of the contagion by public health measures such as masking, social distancing, self isolation and hygiene, or more recently by vaccines^7,8^

There are also pharmaceutical/ therapeutic agents with antiviral properties, being repurposed to urgently treat or serve as chemoprevention for COVID 19. One such drug is Ivermectin, which has exhibited broad spectrum anti-parasitic, anti-bacterial and antiviral properties against many RNA viruses^9,10^. Ivermectin is extensively used, with good safety profile in Nigeria and other African nations in treating ocular onchocerciasis^11^. Of more current import, Ivermectin was shown to exhibit a 5000-fold reduction in SARS-C0V-2 viral RNA in vitro in Vero-h/SLAM cells in a study from Australia.^12^

There are several mechanisms by which Ivermectin may inhibit SARS-CoV-2 in COVID 19 patients, including by inhibition of RNA-dependent RNA polymerase (RdRP) required for viral replication,^13^ abolition of importin-α/β1 heterodimer nuclear transport of SARS-CoV-2 from the cytosol to the nucleus, and inhibition of viral mRNA and viral protein translation^14^.

However, there has been skepticism as to whether the virucidal IC50 of Ivermectin against SARS-CoV-2 of 2.4uM (obtained in vitro) could be feasible or attainable in humans or patients with COVID 19. This is because a 10-fold dose of Ivermectin (120mg) simulations-based kinetics in cattle still did not yield peak drug levels (Cmax) approaching the IC50 for in vitro SARS-CoV-2 inhibition^15^. In yet another simulation, based on human pharmacokinetics of different potential antiviral SARS-CoV-2 repurposed drugs, Ivermectin was one of the drugs predicted to have 10-fold concentrations higher than their reported 50% effective concentration [EC50].^16^ There is thus conflicting report on simulations from cattle and human pharmacokinetics in the effective anti-SARS-CoV-2 concentration attainable by Ivermectin dosing. Ivermectin has a long half life (t^1/2^) of 81-91 hours, and is highly lipophilic with a high volume of distribution (Vd), indicating preferential lung and tissue accumulation.^17^

Pharmacodynamically, Ivermectin dose-dependently inhibits lipopolysacharide (LPS) induced release of inflammatory cytokines (interleukins) in mice and improved LPS-induced survival.^18^ Collectively, there are were multiple pharmacodynamic and pharmacokinetic basis that suggest a potential utility and efficacy of Ivermectin in COVID 19.

We therefore tested the hypothesis that Ivermectin will exert a clinically and therapeutically beneficial effect in mild to moderate COVID 19 patients in a randomized double blind controlled clinical trial in Nigerian COVID 19 patients with RT-PCR proven SARS-CoV-2 positivity.

## METHODOLOGY

The study was a proof of concept (PoC), double blind, randomized controlled trial, of a parallel group, dose-response design. There were 3 treatment groups to which COVID 19 positive patients were randomized. One group received Ivermectin 6mg (given every 84 hours) twice a week, another group received Ivermectin 12mg (given every 84 hours) for 2 weeks, and the third group received lopinavir / ritonavir daily for 2 weeks.

The study protocol received ethical review and approval of the Institutional Review Board of the Lagos University Teaching Hospital (LUTH), Lagos, Nigeria. The protocol was also reviewed and approved by the Nigerian Agency for Food and Drug Administration and Control (NAFDAC) in Lagos.

The patient inclusion criteria were COVID 19 PCR proven positive patients, who gave informed, written consent to participate in the study, and were either asymptomatic or had mild/moderate symptoms.

Study exclusion criteria were COVID 19 negative patients, patients who had COVID pneumonia or requiring ventilator therapy, renal failure, thromboembolic complications, or unconscious by reduced Glasgow Coma Scale.

There were 62 patients randomized to the 3 treatments. COVID 19 PCR testing was undertaken at baseline (pretreatment time 0 hour) and after dosing at 0 hours, 84 hours, 168 hours, 232 hours and 336 hours.

The baseline demographic, clinical, laboratory and virological data of the 3 patient groups is summarized on Table 1.

**Table 1:**
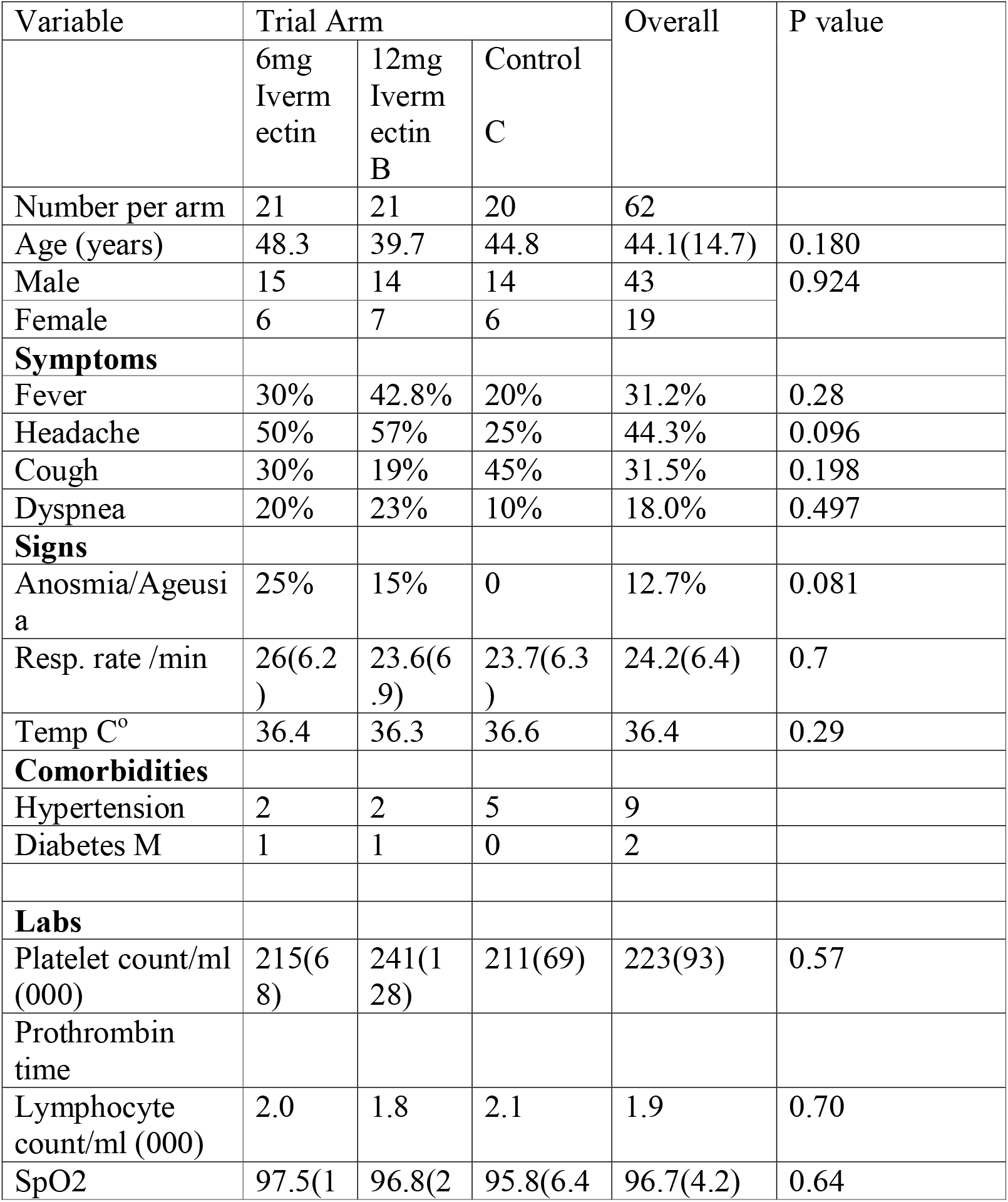
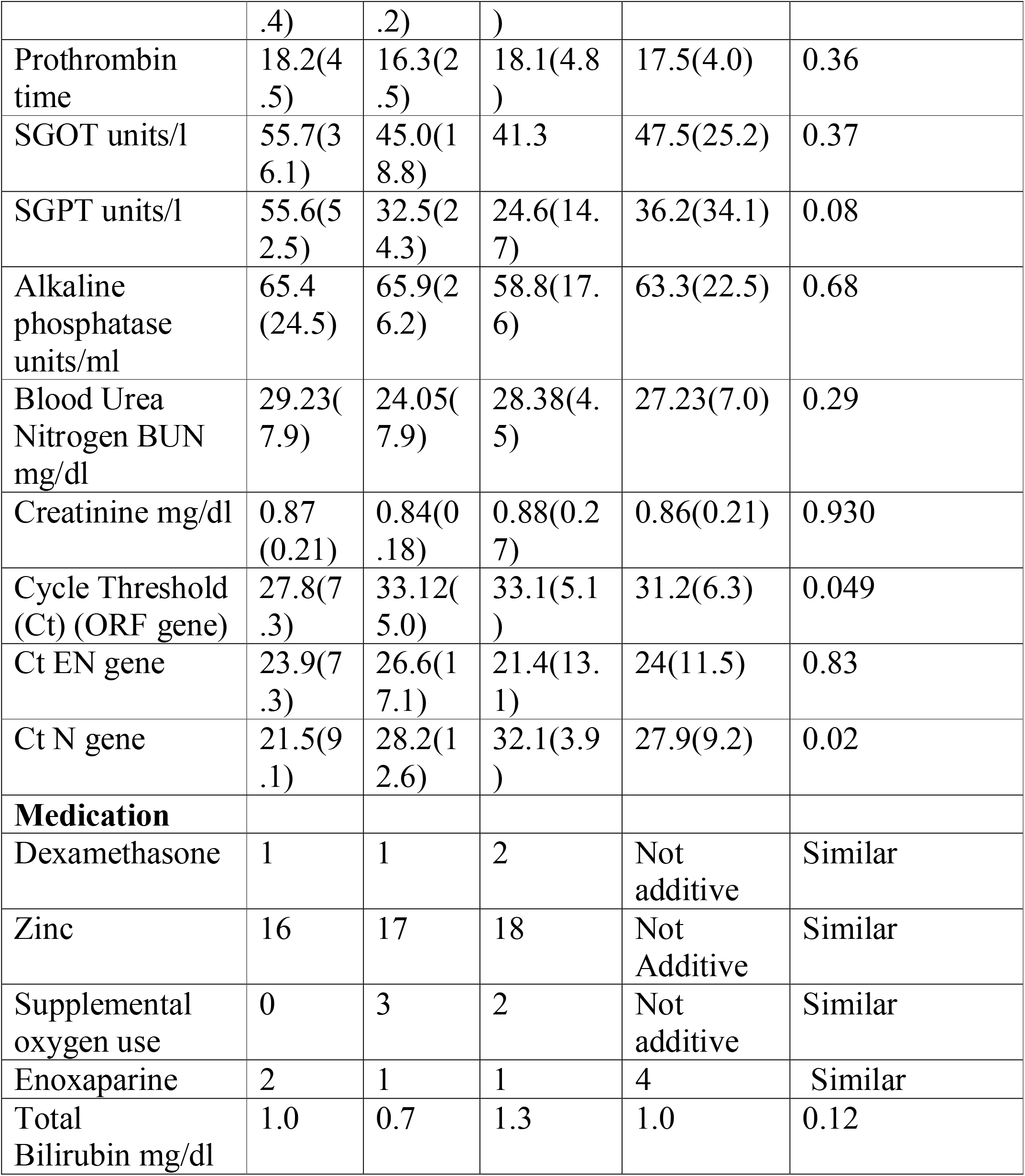
Distribution of baseline Variables by Trial arm (SD/95%CI)

### RT - PCR

COVID 19 test was by RT polymerase chain reaction (PCR) testing for 3 genes. Using a geneexpert machine and testing simultaneously for three genes (Orf, EN, N) A positive COVID 19 test requires all the 3 genes to be present, and a negative test requires all the genes to be functionally absent.

The Cycle threshold (Ct) of the gene tester of more than 40 was regarded as negative, and values below 40 are positive where the Ct value bears an inverse logarithmic relationship to the SARS-CoV-2 viral load.^19,20^

Routine biochemistry, hematology, arterial oxygen saturation (Pa02) temperature and clinical data were gathered, and prognostic ones were recorded at the aforementioned times.

## RESULTS

This study was undertaken between May and November 2020. A general description of the study population and the spread over three arms is found in Table 1. Sixty-three patients with positive PCR result were randomised into three arms of the study. There was one withdrawal, thus sixty-two patients completed the study. The average age was 44.1years (SD14.7), ranging from 20-82. There were 43 males and 19 females. The patients had mild to moderate clinical symptoms and none of them required ventilator, although five required intranasal oxygen, 3 in the 12mg arm (B) and two in the control arm (C). One third of the patients reported with a fever and cough, while 44% and 18% respectively reported with headache and difficulties with breathing. 12% reported with anosmia/ageusia. The commonest comorbidities were Diabetes Mellitus (DM) (2) and Hypertension (9), while some had combined hypertension and DM. Some patients required concomitant medications such as dexamethasone, enoxaparin, and supplemental oxygen.

The effectiveness of randomization was assessed, and the results are displayed in Table 1. In all, twenty-one patients were each randomised into the 6mg (A) and 12mg (B) Ivermectin arms while twenty went into the control arm (C).

There was no significant difference in the distribution of the age, sex and symptoms, comorbidities, blood counts, prothrombin time, liver function and kidney function tests. There were however slight differences in the baseline Cycle threshold (Ct) values, being lower in the A arm than the other two arms with regards to the ORF and N genes, but similar for the EN gene. The distribution of other supplemental medications taken by participants, aside from Ivermectin, was broadly similar. These included Zinc, ascorbic acid, vitamin D and Azithromycin.

### Changes in variables over time was assessed

The time to SARS-CoV-2 negativity is described in Figures 1A and B and Table 2. Ivermectin significantly and dose-dependently reduced time to negativity compared to the control group. The mean days-to-negative in the control arm was 9.15 (CI 5.68 to 12.62) while for combined Ivermectin arms it was 5.33 (CI 4.33 to 6.32). Average days-to-negative in the combined Ivermectin arms (any Ivermectin) was thus shorter by 3.83 days (95% CI 6.54 to 1.11) compared to controls P=0.0066. (Student t-test.)

**Table 2:** Days to negative in the three trial arms.

| Trial Arm | Mean days-to- negative<br>(95% Confidence Interval) | Standard deviation | N |
| --- | --- | --- | --- |
| A<br>6mg Ivermectin | 6.0<br>(4.61 - 7.38) | 2.96 | 20 |
| B<br>12mg Ivermectin | 4.65<br>(CI 3.15- 6.15) | 3.2 | 20 |
| A&B<br>(Any Ivermectin) | 5.33<br>(CI 4.33-6.32) | 3.12 | 40 |
| C<br>Control | 9.15<br>(CI 5.68- 12.62) | 7.42 | 20 |
| Total study<br>population | 6.6<br>(CI 5.25-7.95) | 5.24 | 60 |

Tests were not completed in two cases.
The one-way ANOVA $P > F = 0.017$ .
T-test comparing “Any Ivermectin” and control $P = 0.0066$

**Figure 1A:**
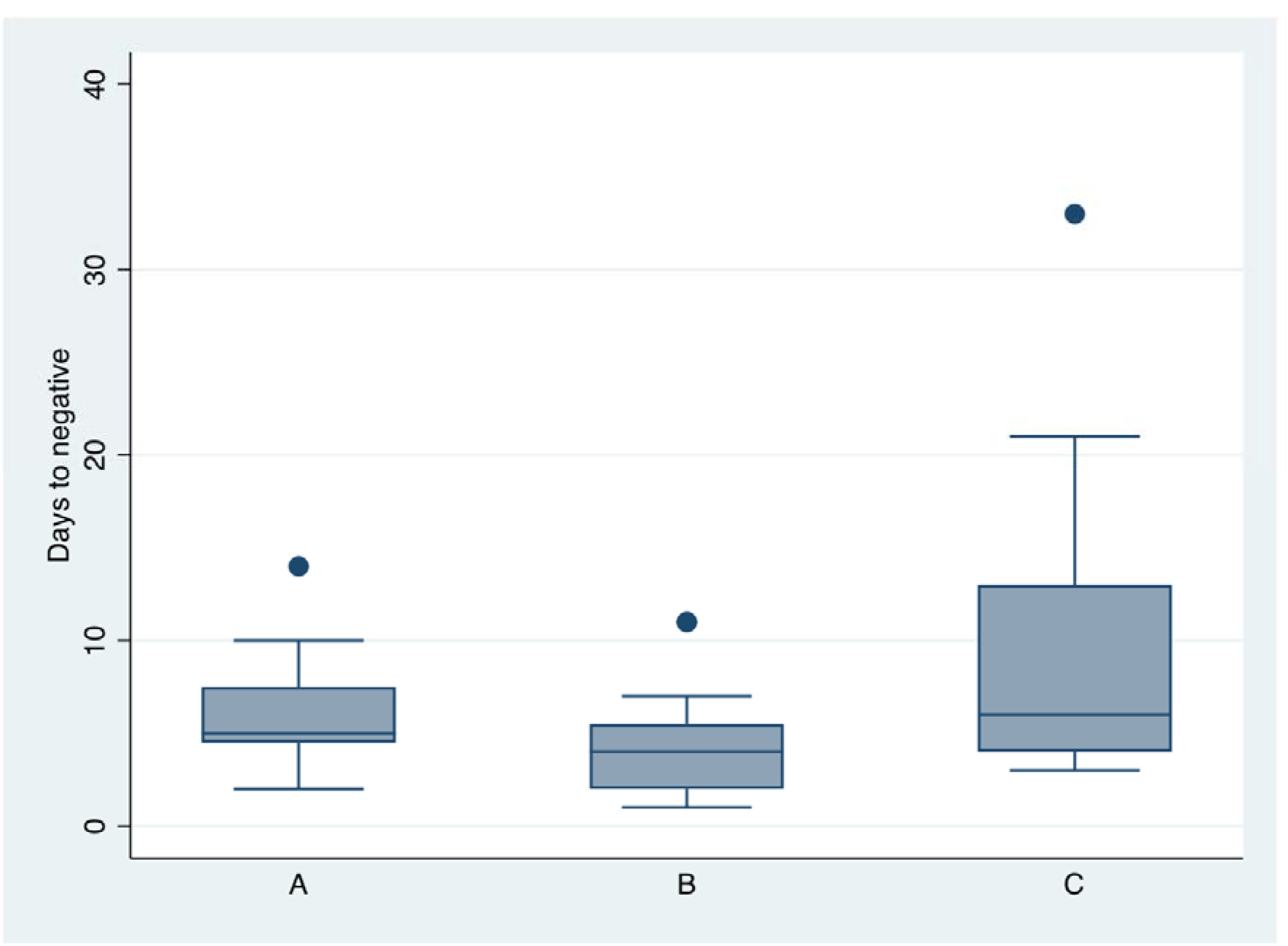
Boxplot showing distribution of Days to Negative between the three arms of the study: A=6mg Ivermectin, B=12mg Ivermectin C=Control.

**Figure 1B:**
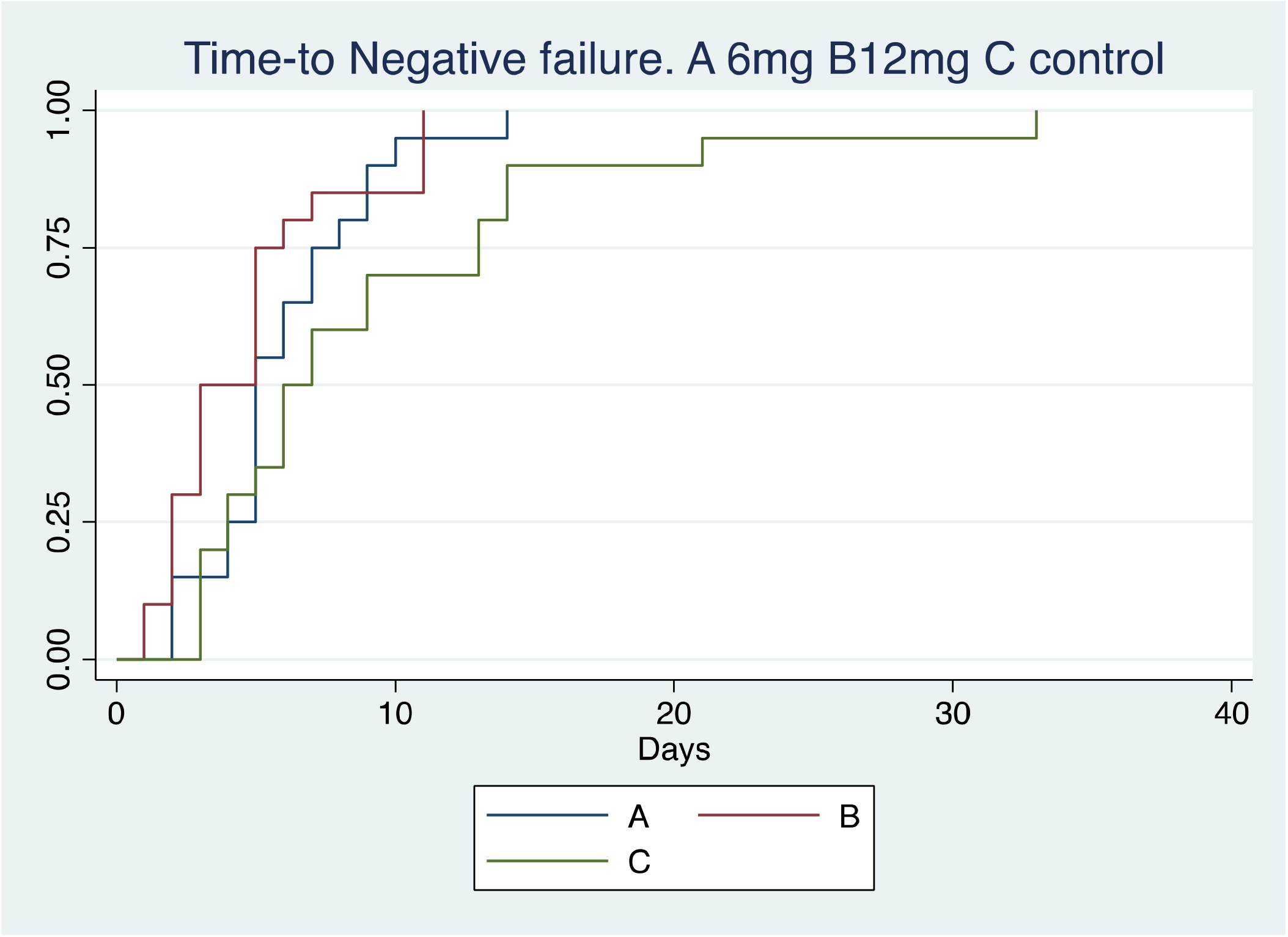
Kaplan Meir Curve showing time sequence of days to negativity in the three arms of the study. Log Rank P>Chi= 0.036. Hazard Ratio for 6mg arm 1.68, (P=0.120, CI 0.87 to 3.25) Hazard Ratio for 12mg arm 2.38 (P=0.011, CI 1.22 to 4.65)

Mean days-to-negative for the 12mg arm was however shortened by 4.5 days, and by 3.15 days for the 6mg arm compared to controls. These differences were significant by ANOVA P>F =0.0179.

The distribution of the days-to-negative are depicted in Figure 1A. Figure 1B depicts a Kaplan Meier (KM) curve comparing time to negative (failure) for the three arms. Log rank test for equality of survival functions gave P>chi = 0.0363. Figure 1C depicts KM curve comparing ‘Any Ivermectin’ with controls. P>chi=0.019.

**Figure 1C:**
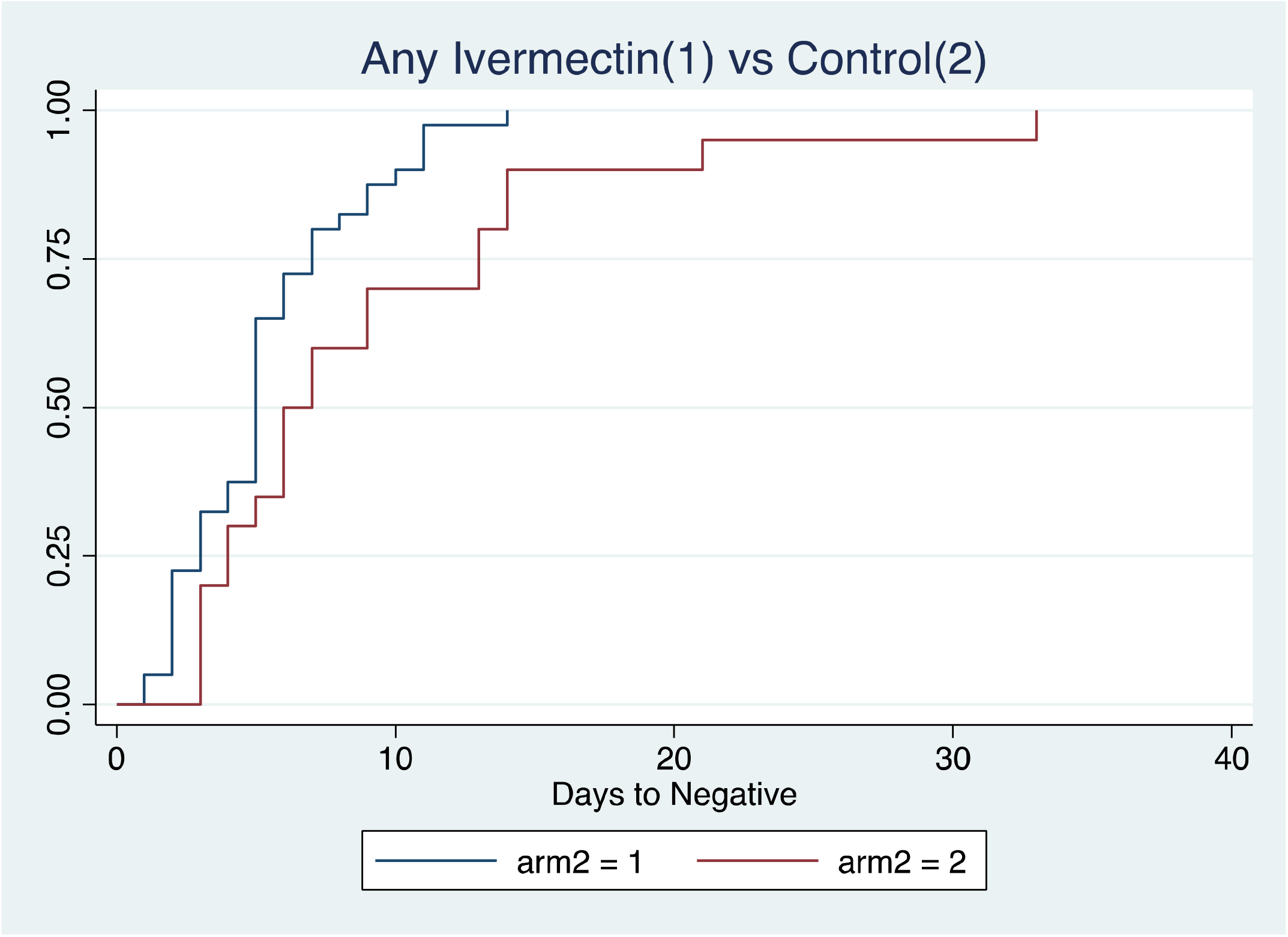
Kaplan Meir Curve showing time sequence of days-to-negativity, ‘Any Ivermectin’ versus Control. Log Rank P>Chi= 0.019. Hazard Ratio 1.96, P= 0.024 CI 1.09 −3.51

Cox proportional hazard model enabled computation of Hazard Ratio (HR) using the control arm as base. HR for the 6mg arm was 1.68, (P=0.120, CI 0.87 - 3.25) while for the 12mg arm, HR was 2.38 (P=0.011, CI 1.22 - 4.65). This suggests that the 12 mg treatment arm will cause the patient to progress 2.38 times faster than the control group. Although there is a HR of 1.68 for the 6mg arm, it did not achieve statistical significance.

The HR for “Any Ivermectin” was 1.96, P= 0.024, CI 1.09 −3.51.

A 2-way 2 factor Repeated Measure Analysis of Variance (RAMOVA) concerning SARS-CoV-2 PCR assays was carried out to compare means and variances of each of the three treatments at the different time points to obtain treatment effect, Time effect and Time Treatment interaction. A plot of the Means and Standard Error of the Means of 20 patients in each of group A, B, C is shown in Figure 2. There was a significant treatment (p=0.035) and time effect (p <0.0001) of Ivermectin on COVID 19 ranked scores compared to controls.

**Figure 2:**
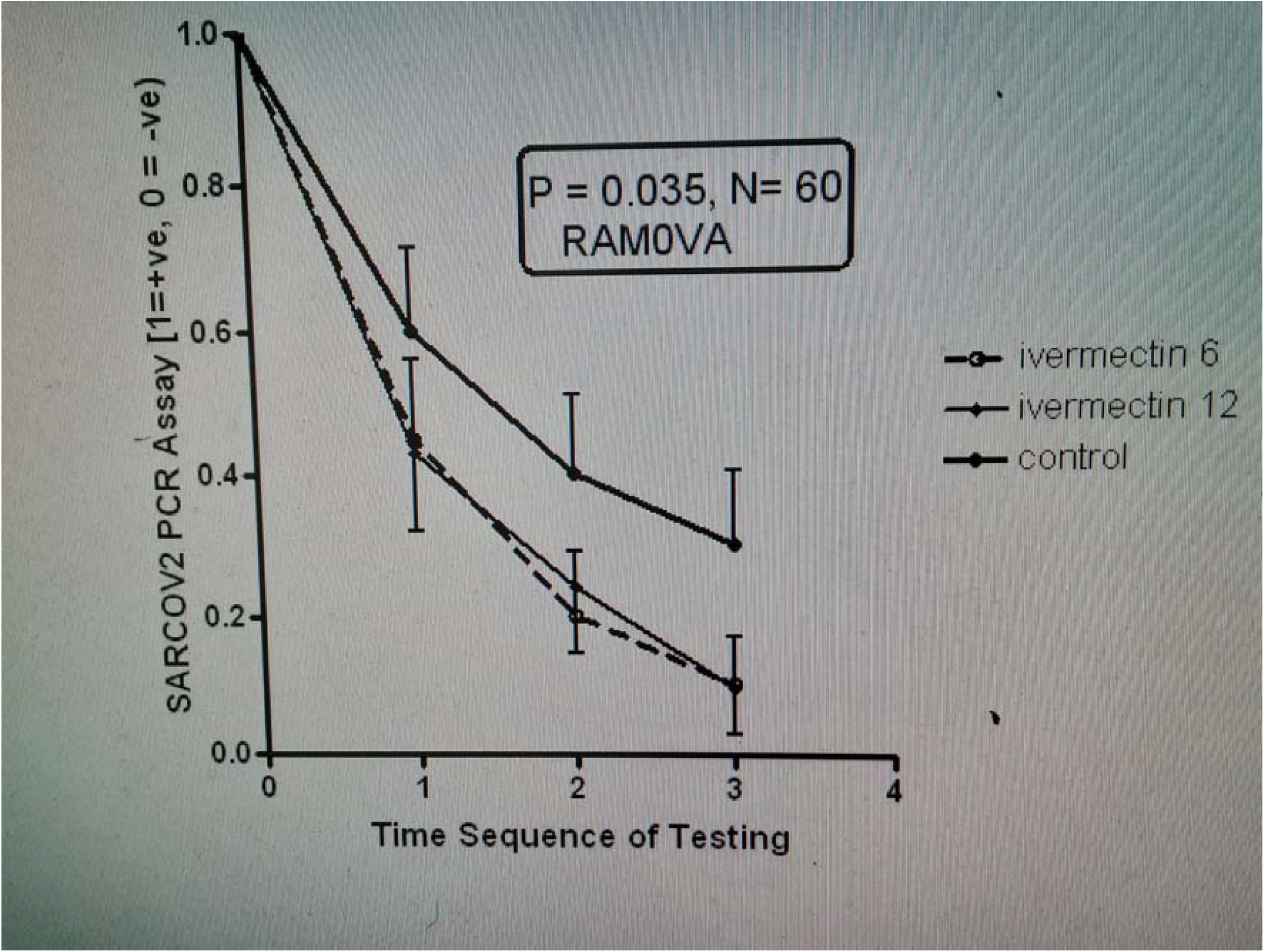
2-Way Repeat Measures of Analysis of Variance (RAMOVA) SARSCOV2 PCR ASSAY. Key: Time sequence 1=+84hrs 2=+168hrs 3=+232hrs 4=+336hrs

The likelihood of negativity by day five was explored. The Ivermectin arm was 3.45 times more likely to go negative by or before day five, P=0.0271, 95% CI 1.12-10.63. This effect is slightly mitigated by sex (adjusted OR 3.44 P>z 0.031 CI= 1.12 −10.6) but more significantly by age (OR 2.77 P>z 0.113 CI= 0.79 to 9.8).

Changes in clinical and laboratory parameters at baseline and at seven days (or as otherwise stated) were observed for the three arms and recorded in Table 3. Day seven was used as a midway point in the trial. Of note was that there was a moderate increase in SpO2 in the Ivermectin arm, although this did not attain significance (P=0.098). The difference from baseline to highest attained SpO2 during the study for each participant is also depicted in Figure 3A, in which baseline figures are compared with highest attained SpO2. (P=0.073)

**Table 3:**
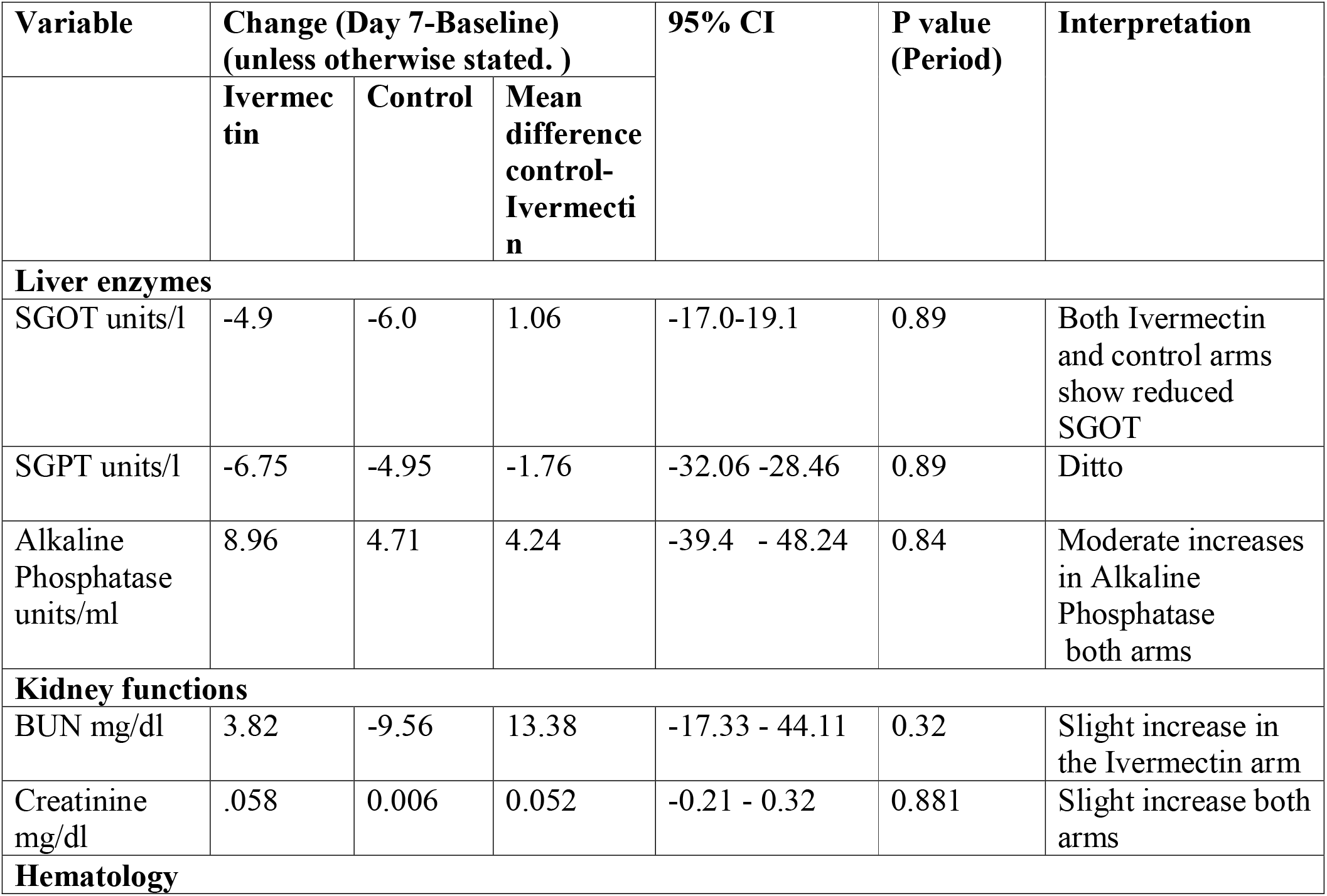
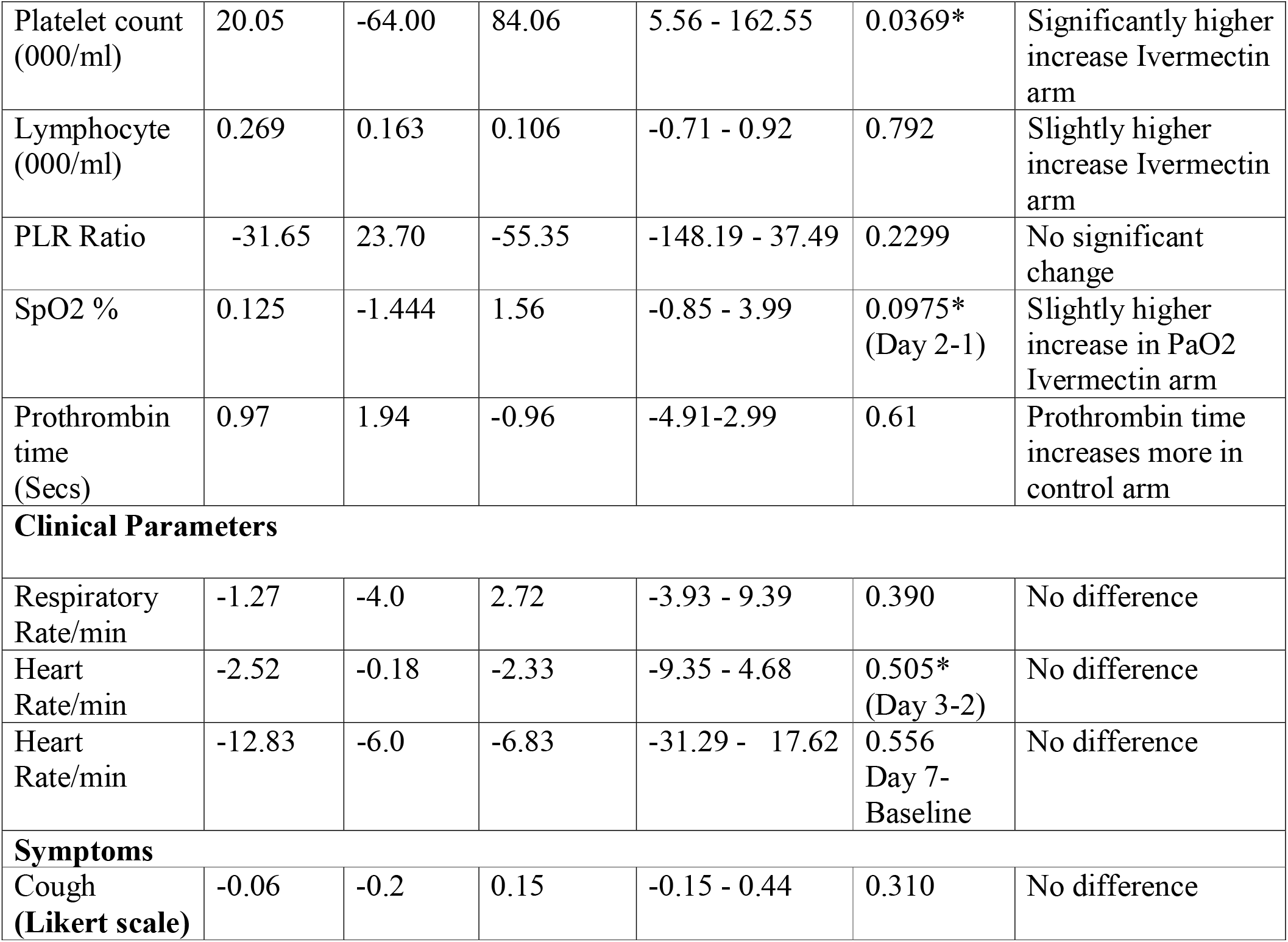
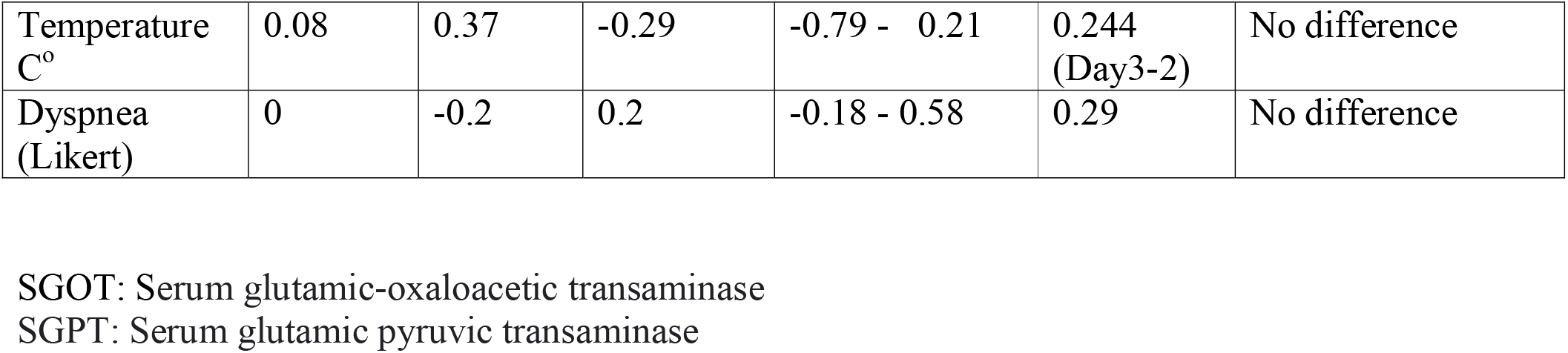
Changes in variables over time, Ivermectin versus Control arm.

**Figure 3A:**
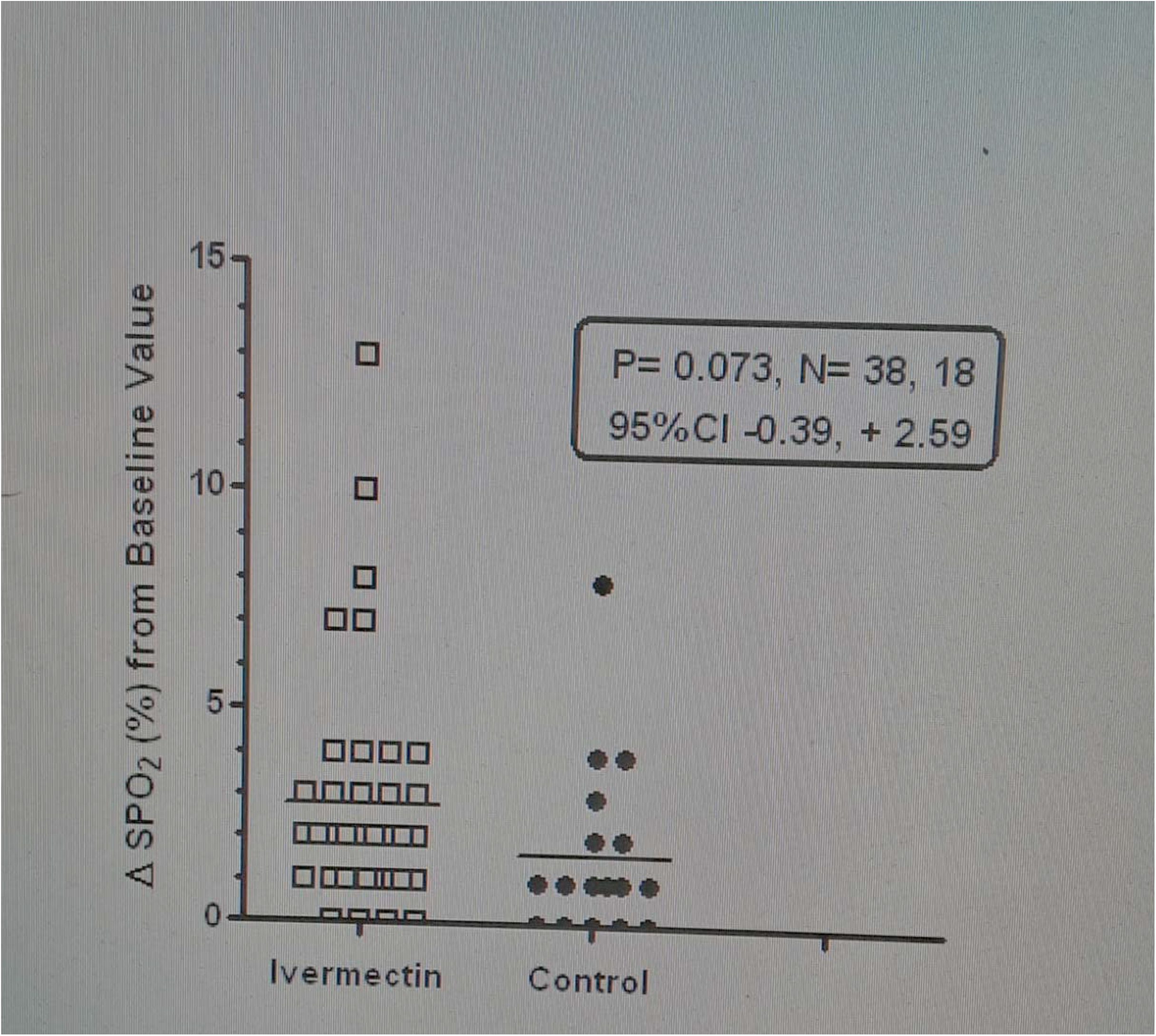
Difference in change in SpO2 from baseline to highest attained in study.

Because the drug Ivermectin is essentially metabolized in the liver via CYP34A, changes in liver enzymes such as SGOT (serum glutamic-oxaloacetic transaminase,) SGPT (Serum glutamic pyruvic transaminase) and Alkaline Phosphatase were analysed. Both arms of treatment showed reduction in SGOT and SGPT and moderate increases in Alkaline phosphatase. None of these were statistically significant.

There were also no significant differences in changes in kidney function tests such as Blood Urea Nitrogen and Creatinine between the three groups.

There was a notable significant increase in platelet counts in the ivermectin arm relative to the control arm (P=0.037). See Figure 3B. There was also a moderate but not significant relative increase in lymphocyte count. The overall Platelet Lymphocyte Ratio (PLR) was not significantly changed (Table 2). In Figure 3C, we note a statistically significant (P=0.0055) negative correlation between days-to-negative and increase in platelet count. The higher the change in platelet count, the fewer the days-to-negative. Pearson’s r= −0.53, R^2^=0.28

**Figure 3B:**
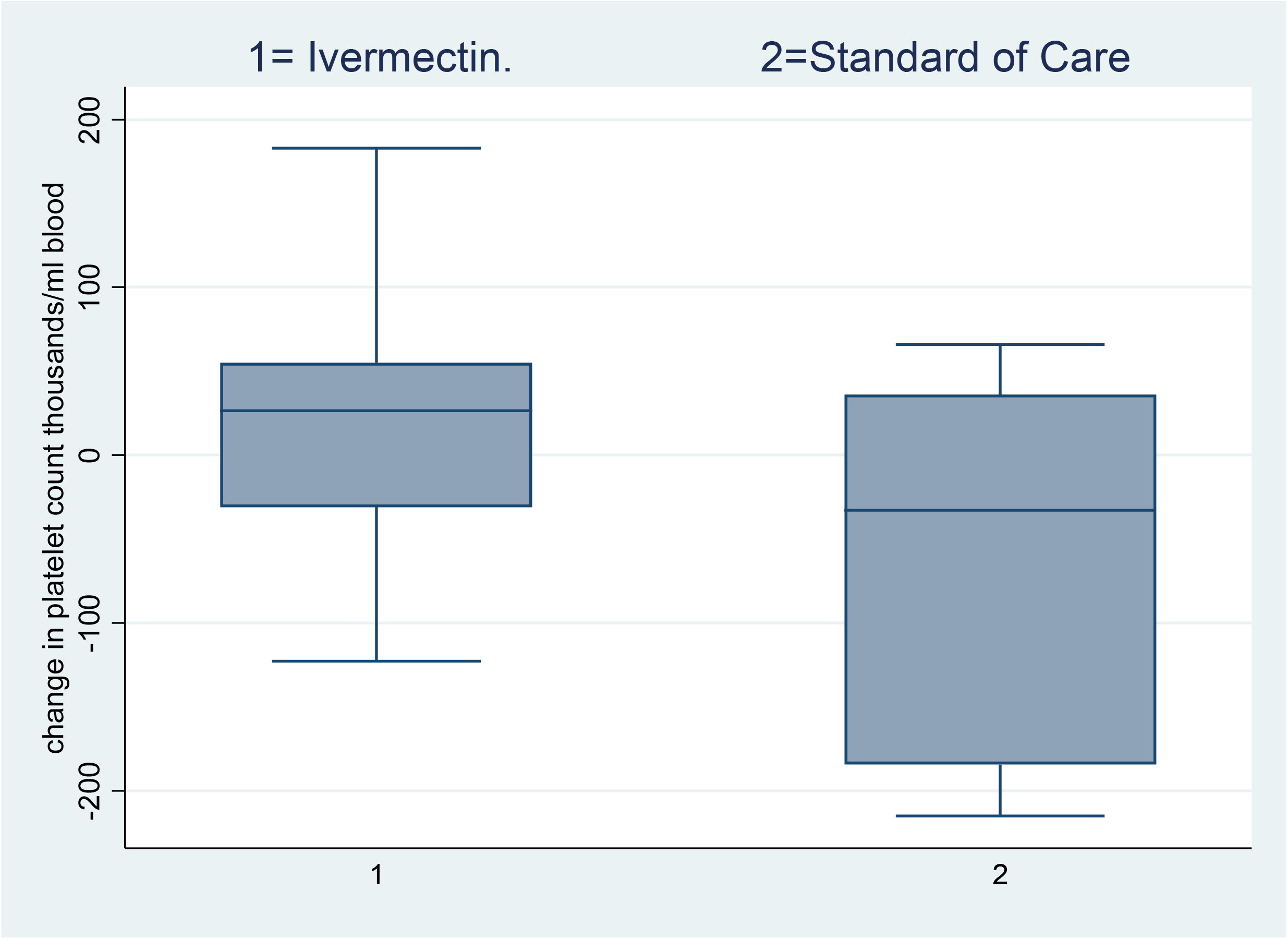
Change in platelet count (Day7-Baseline) Ivermectin group versus control (Standard of Care) Mean difference = 84.05556 95% CI =5.56 - 162.55, P= 0.0369.

**Figure 3C:**
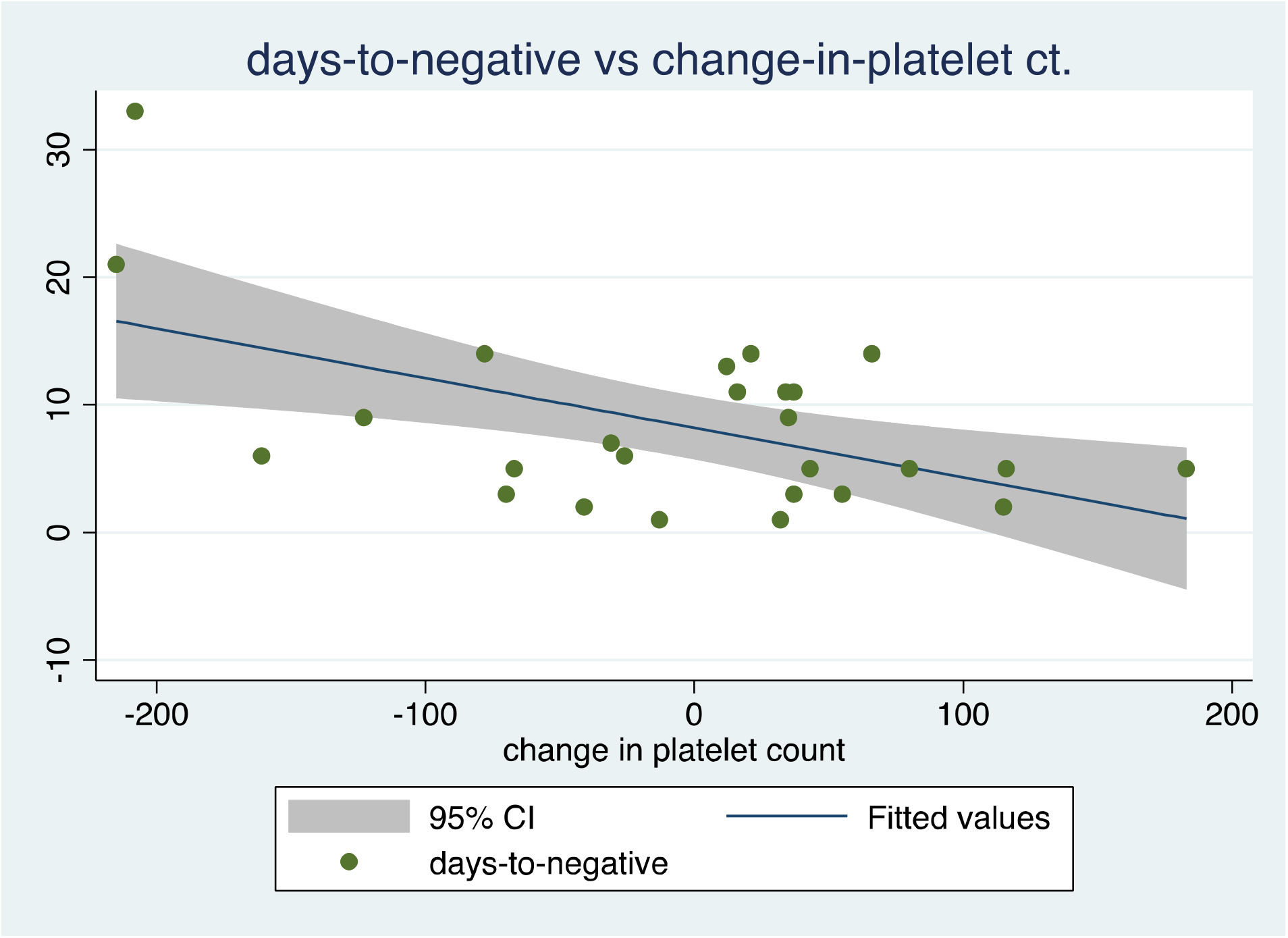
Scatterplot exploring relationship between change in platelet count and Days-to-negative. PCR. r= −0.53, R^2^=0.28 P>F=0.0055.

There were slight increases in prothrombin time across all arms, more so in the control arm. Rate of increase was not significantly different across arms.

There were no significant changes over time, across the three groups in clinical parameters such as Respiratory rate, Heart rate, Temperature, and symptoms such as cough and dyspnea as assessed by Likert scales.

There was no significant overall effect of age on days-to-negative by linear regression analysis (r =0.046 p = 0.728). However, those in the 30-40-year age band had a lower time to negative relative to others.

No adverse effects of Ivermectin was reported in response to questioning or spontaneous report.

Symptomatic improvement was seen in all patients, with resolution of fever dypnea and other signs. There was no mortality and the patients remained well on follow up.

## DISCUSSION

Our findings show a statistically significant and dose dependent effect of Ivermectin to reduce the time to SARS-CoV-2 negativity in RT-PCR COVID 19 positive patients (p =0.0066) and significant treatment (p=0.035) and time effect (p <0.0001) of Ivermectin on COVID 19 ranked scores compared to controls by RAMOVA. A Hazard Ratio of 2.38 in the 12mg arm suggests that negativity events such as described above are likely to occur more than twice as fast with Ivermectin. In addition, Ivermectin appears to be 3.45 times more likely (P=0.0271, 95% CI 1.12-10.63.) to induce negativity by day 5 compared to controls. Collectively, these results demonstrate a likely beneficial treatment effect of Ivermectin, to reduce the duration of illness, elicit faster recovery and diminution of qualitative indices of SARS-CoV-2 viral load compared to the usual treatment.

These results are consistent with those of Ahmed et al^21^ who demonstrated a significant reduction in time to COVID 19 virological clearance with Ivermectin of 3 days on average, albeit at a higher dose of 12mg daily for 5 days in Bangladesh. Similar findings were reported by Elgazzar and colleagues in Egypt^22^.

Our results demonstrate a dose-response in the effects of Ivermectin 6mg and 12mg twice weekly in time to SARS-CoV-2 negativity. The time (mean +/− SD) to negativity (days) in the treatments were; 6mg; 6+/−2.95, 12 mg; 4.65 +/− 3.19, and the controls 9.15+/− 7.26, p =0.02 ANOVA. When both Ivermectin groups were combined (with a new mean 5.34 +/− 0.07 days n=41) the mean difference SEM from placebo control, of - 3.81 +/− 1.34 days was statistically significant p = 0.0066, 95% CI −1.13 to - 6.49 days.

These collectively suggest that the 12 mg dose achieved a faster SARS-CoV-2 virological clearance compared to the 6 mg dose, and that using these standard doses of Ivermectin employed in onchocerciasis treatment caused a 3.8 days faster virological clearance, and clinical amelioration compared to controls on lopinavir/ritonavir.

The findings also provide Proof of concept (Poc) of Ivermectin efficacy against SARS-CoV-2 at the doses that were initially considered inadequate based on pharmacokinetic simulations^15,17^

The progressive reduction in COVID 19 positivity in the Ivermectin-treated group over time, as well as the treatment effect were sustained with no time-treatment interactions (Figure 2. Table 2). This was associated both with symptomatic improvements as well as Ivermectin-induced reversal of abnormal COVID 19 prognostic parameters. Ivermectin treatment was associated with a strong trend to an increase in arterial oxygen saturation (SPO2%) compared to controls (See Figure 3A) p= 0,073 and 95% CI of −0.39 to 2.59. Pulse oximetry at the finger digits, has recently been shown to exhibit racial divergences with a higher likelihood of overrating SPO2% and leading to “occult hypoxemia” occurring more in black people in comparison to whites^23^ thus, the real change in SPO2 with Ivermectin may have been masked.

Platelet count is another prognostic index in COVID 19 with thrombocytopenia reflecting platelet consumption in SARS-CoV-2 as part of sepsis-induced coagulopathy, even before frank Disseminated Intravascular Coagulation (DIC) manifests as hemorrhage or thrombosis^24^. Ivermectin treatments (combined 6mg and 12 mg dose) caused an increase in platelet count relative to the control group patients with a 95% CI for the difference of 5.55 to 162.55 × 10^9^/L p=0.037 ANOVA, F =4,88, df −24. (See Table 3). Further, there was an inverse correlation (r = - 0. 52, R^2^ = 0. 28, p = 0.005) between the Ivermectin induced increase in platelets and the time to SARS-CoV-2 negativity in the patients (See Figure 3C). These platelet results indicate an Ivermectin effect to reverse a negative prognostic factor, and the increase in platelets is associated with a faster resolution and inhibition of SARS-CoV-2 virological proliferation.

The baseline prothrombin time appeared slightly prolonged in all the patients (>0.15 seconds), but no effect of treatment in any group was apparent. The concurrent trend to Ivermectin improvement of COVID 19-associated arterial hypoxemia, concomitant with a reduction in prothrombotic hematological coagulopathy (reversal of platelet consumption) may be a consequence of ivermectin anti-inflammatory reduction in COVID-19 hypercytokinemia (“cytokine storm’), and is entirely consistent with reduction in mortality and especially in patients with SARS-CoV-2 pulmonary complications.^25-28^ Meta analysis revealed that IL-6 and associated cytokines are correlated with SARS-CoV-2 severity in patients^29^ which are under Janus Kinase (JAK-STAT) signaling pathway.^30^ Given the known effect of Ivermectin to inhibit IL-6, TNF-alpha in vivo and in vitro, and its suppression of NF-kappa B translocation in mice,^18^ a formal controlled study of the effect of Ivermectin on hyper-cytokinemia is now indicated in COVID 19.

There were no significant changes in hepatic and renal functions given that Ivermectin is hepatically metabolized. The gender of participants did not exert any effects on the pharmacodynamic effects of Ivermectin in time to COVID 19 negativity. However, the likelihood of negativity by day 5 was mitigated by age from 3.45 to 2.77.

Ivermectin was remarkably well tolerated and there was no adverse drug event reported spontaneously or in response to inquiry.

In conclusion, Ivermectin exhibited a dose-dependent significant inhibitory effect on SARS-CoV-2. This study provides support for the translation of the in vitro findings of Caly et al^12^ at doses that were initially thought to be suboptimal in humans. The 12mg twice weekly regime appears to confer a superior efficacy. Ivermectin modified prognostic factors such as arterial oxygenation and platelet and hematological indices of SARS-CoV-2 infections. The mechanisms of the benefits were not established in this study, but an effect on JAK-STAT and NF-kappa B related cytokines should be studied in patients.

Further study of Ivermectin 12mg in the pre-exposure prophylaxis and prevention of community transmission of SARS-CoV-2, such as spoke-and-wheel or targeted hotspots treatment, is now warranted, especially as an interim measure in countries that cannot immediately roll out vaccination programs.

## Data Availability

All data referred to in the manuscript is available

https://www.ivercovid.com

## Acknowledgment

**Professor Amam C. Nbakwem**. Professor of Cardiology, Department of Medicine, College of medicine, University of Lagos.

## References

1. Huang C, Wang Y, Li X, Ren L, Zhao J, Hu Y, et al. Clinical features of patients infected with 2019 novel coronavirus in Wuhan, China. The lancet. 2020;395(10223):497–506.

2. Cucinotta D, Vanelli M. WHO Declares COVID-19 a Pandemic. Acta Bio Medica Atenei Parm. 2020;91(1):157–60.

3. Elfein J. https://www.statista.com/statistics/1103046/new-coronavirus-COVID19-cases-number-worldwide-by-day-GoogleSearch.

4. Carfì A, Bernabei R, Landi F, for the Gemelli Against COVID-19 Post-Acute Care Study Group. Persistent Symptoms in Patients After Acute COVID-19. JAMA. 2020 Aug 11;324(6):603.

5. Khullar D, Bond AM, Schpero WL. COVID-19 and the Financial Health of US Hospitals. Jama. 2020;323(21):2127–8.

6. Jackson JK, Weiss MA, Schwarzenberg AB, Nelson RM, Sutter KM, Sutherland MD. Global Economic Effects of COVID-19. :132.

7. Polack FP, Thomas SJ, Kitchin N, Absalon J, Gurtman A, Lockhart S, et al. Safety and Efficacy of the BNT162b2 mRNA COVID-19 Vaccine. N Engl J Med. 2020 Dec 31;383(27):2603–15.

8. Anderson EJ, Rouphael NG, Widge AT, Jackson LA, Roberts PC, Makhene M, et al. Safety and Immunogenicity of SARS-CoV-2 mRNA-1273 Vaccine in Older Adults. N Engl J Med. 2020 Dec 17;383(25):2427–38.

9. Heidary F, Gharebaghi R. Ivermectin: a systematic review from antiviral effects to COVID-19 complementary regimen. J Antibiot (Tokyo). 2020 Sep;73(9):593–602.

10. Jans DA, Wagstaff KM. The broad spectrum host-directed agent ivermectin as an antiviral for SARS-CoV-2L? Biochem Biophys Res Commun [Internet]. 2020 Oct 21 [cited 2021 Jan 1]; Available from: http://www.sciencedirect.com/science/article/pii/S0006291X20319598

11. Abiose A, Jones BR, Murdoch I, Cassels-Brown A, Babalola OE, Alexander ND, Evans J, Ibrahim UF, Mahmood AO, Cousens SN, Nuhu I. Reduction in incidence of optic nerve disease with annual ivermectin to control onchocerciasis. The Lancet. 1993 Jan 16;341(8838):130–4.

12. Caly L, Druce JD, Catton MG, Jans DA, Wagstaff KM. The FDA-approved drug ivermectin inhibits the replication of SARS-CoV-2 in vitro. Antiviral Res. 2020 Jun 1;178:104787.

13. Parvez MSA, Karim MA, Hasan M, Jaman J, Karim Z, Tahsin T, et al. Prediction of potential inhibitors for RNA-dependent RNA polymerase of SARS-CoV-2 using comprehensive drug repurposing and molecular docking approach. 2020 Apr 15 [cited 2021 Jan 1]; Available from: https://arxiv.org/abs/2004.07086v1

14. Yang SNY, Atkinson SC, Wang C, Lee A, Bogoyevitch MA, Borg NA, et al. The broad spectrum antiviral ivermectin targets the host nuclear transport importin α/β1 heterodimer. Antiviral Res. 2020 May 1;177:104760.

15. Schmith VD, Zhou J (Jessie), Lohmer LRL. The Approved Dose of Ivermectin Alone is not the Ideal Dose for the Treatment of COVID-19. Clin Pharmacol Ther. 2020;108(4):762–5.

16. Arshad U, Pertinez H, Box H, Tatham L, Rajoli RK, Curley P, et al. Prioritisation of Anti-SARS-Cov-2 Drug Repurposing Opportunities Based on Plasma and Target Site Concentrations Derived from their Established Human Pharmacokinetics. Clin Pharmacol Ther. 2020;

17. Muñoz J, Ballester MR, Antonijoan RM, Gich I, Rodríguez M, Colli E, et al. Safety and pharmacokinetic profile of fixed-dose ivermectin with an innovative 18mg Tablet in healthy adult volunteers. PLoS Negl Trop Dis. 2018 Jan 18;12(1):e0006020.

18. Zhang X, Song Y, Ci X, An N, Ju Y, Li H, et al. Ivermectin inhibits LPS-induced production of inflammatory cytokines and improves LPS-induced survival in mice. Inflamm Res. 2008 Nov 1;57(11):524–9.

19. Nandini S, Sundararaj SJ, Akihide R. Interpreting Diagnostic Tests for SARS-CoV-2 Au JAMA. Publ Online May. 2020;6.

20. James J. Dunn. Laboratory Diagnostics and Testing Guidance for COVID-19: Laboratory Studies, Specimen Selection, Collection, and Transport, Nucleic Acid Detection [Internet]. [cited 2021 Jan 2]. Available from: https://emedicine.medscape.com/article/2500138-overview

21. Ahmed S, Karim MM, Ross AG, Hossain MS, Clemens JD, Sumiya MK, et al. A five day course of ivermectin for the treatment of COVID-19 may reduce the duration of illness. Int J Infect Dis [Internet]. 2020 Dec 2 [cited 2021 Jan 1]; Available from: http://www.sciencedirect.com/science/article/pii/S1201971220325066

22. Elgazzar A, Hany B, Youssef SA, Hafez M, Moussa H. Efficacy and Safety of Ivermectin for Treatment and prophylaxis of COVID-19 Pandemic. 2020;

23. Sjoding MW, Dickson RP, Iwashyna TJ, Gay SE, Valley TS. Racial Bias in Pulse Oximetry Measurement. N Engl J Med. 2020 Dec 17;383(25):2477–8.

24. Liao D, Zhou F, Luo L, Xu M, Wang H, Xia J, et al. Haematological characteristics and risk factors in the classification and prognosis evaluation of COVID-19: a retrospective cohort study. Lancet Haematol. 2020 Sep 1;7(9):e671–8.

25. Rajter JC, Sherman MS, Fatteh N, Vogel F, Sacks J, Rajter J-J. Use of Ivermectin Is Associated With Lower Mortality in Hospitalized Patients With Coronavirus Disease 2019: The ICON Study. Chest [Internet]. 2020 Oct 13 [cited 2021 Jan 1]; Available from: http://www.sciencedirect.com/science/article/pii/S0012369220348984

26. Moore JB, June CH. Cytokine release syndrome in severe COVID-19. Science. 2020;368(6490):473–4.

27. DiNicolantonio JJ, Barroso-Arranda J, McCarty M. Ivermectin may be a clinically useful anti-inflammatory agent for late-stage COVID-19. Open Heart. 2020 Sep 1;7(2):e001350.

28. Hussman JP. Cellular and Molecular Pathways of COVID-19 and Potential Points of Therapeutic Intervention. Front Pharmacol [Internet]. 2020 [cited 2021 Jan 2];11. Available from: https://www.frontiersin.org/articles/10.3389/fphar.2020.01169/full

29. Mojtabavi H, Saghazadeh A, Rezaei N. Interleukin-6 and severe COVID-19: a systematic review and meta-analysis. Eur Cytokine Netw. 2020 Jun 1;31(2):44–9.

30. Luo W, Li Y-X, Jiang L-J, Chen Q, Wang T, Ye D-W. Targeting JAK-STAT Signaling to Control Cytokine Release Syndrome in COVID-19. Trends Pharmacol Sci. 2020 Aug 1;41(8):531–43.

